# Distinct subgroups in gastroparesis defined by simultaneous body surface gastric mapping and gastric emptying breath testing

**DOI:** 10.1101/2024.11.21.24317043

**Authors:** Chris Varghese, I-Hsuan Huang, Gabriel Schamberg, Stefan Calder, Christopher N. Andrews, Greg O’Grady, Jan Tack, Armen A Gharibans

## Abstract

**Background:** Gastroparesis is a heterogeneous disorder with several contributing pathophysiologies. In this study we used simultaneous body surface gastric mapping (BSGM) and gastric emptying breath testing (GEBT) to subgroup patients with gastroparesis based on dynamic spectral meal response profiles and emptying rate.

**Methods:** Patients with chronic gastroduodenal symptoms and negative gastroscopy underwent simultaneous BSGM and gastric emptying breath test (GEBT) with 30 minutes fasting and 4 hours postprandial recording. In addition to standard metrics, the BSGM ‘Meal Response Ratio’ (MRR) compared amplitude in the first 2 hours postprandially to the subsequent 2 hours (lagged meal response ≤1).

**Results:** 143 patients underwent simultaneous BSGM and GEBT (79% female, median age 31 years, median BMI 23 kg/m^2^). Delayed emptying occurred in 25.2% (n = 36). Those with a lagged meal response had longer T_1/2_ (median 95.0 [IQR 59-373] vs median 78.0 [IQR 31-288], p=0.009) and higher rates of delayed emptying (42.9% vs 16.7% p = 0.03). BSGM phenotypes identified in patients with delayed emptying were: lagged meal response (25%), low gastric amplitude / rhythm stability (30.6%), elevated gastric frequencies (11.1%), and normal BSGM spectral analysis (33.3%). T_1/2_ weakly correlated with worse total symptom burden score (r = 0.18, p = 0.03).

**Conclusion:** Combined BSGM and gastric emptying testing defines subgroups of gastroparesis based on contributing disease mechanisms, including a novel group with delayed post-prandial onset of gastric motor activity. Improved patient phenotyping in gastroparesis may enable improved therapeutic targeting through these biomarkers of disease processes.

## Introduction

Gastroparesis is defined on the basis of delayed gastric emptying in the absence of mechanical obstruction, with characteristic symptoms of nausea, vomiting, postprandial fullness, early satiety.^1^ Up to 1.8% of the population have symptoms characteristic of gastroparesis although fewer than 0.2% are diagnosed with confirmatory transit testing.^2^ Defining and managing gastroparesis remains challenging owing to labile gastric emptying results,^3^ poor correlations with symptoms,^4^ and overlap with functional dyspepsia and chronic nausea and vomiting syndromes.^3^

Gastric emptying breath testing (GEBT) is an alternative to scintigraphic assessment that avoids radiation exposure, and has the capacity to be done outside of specialist centers. Body surface gastric mapping (BSGM) using the Gastric Alimetry^®^ system (Alimetry, New Zealand), is a non-invasive test of gastric function incorporating high-resolution electrophysiology^5,6 and^ symptom profiles,^7^ which offers complementary information to transit testing.^8^

The clinical utility of confirming the degree of gastric emptying delay in gastroparesis is controversial,^9^ and defining more specific underlying mechanisms for delayed transit through BSGM has been proposed to enhance diagnostic clarity.^8,10^ A multimodal assessment involving an expanded set of physiological biomarkers from both tests may be advantageous in order to better target care towards specific disease mechanisms, while also enabling more specificity in clinical trial enrolment. In this study we therefore applied simultaneous BSGM (Gastric Alimetry) and GEBT to define and evaluate distinct phenotypes of patients with gastroparesis.

## Methods

This was a prospective observational cohort study conducted in Leuven, Belgium (Ethical approvals: S65541). All patients provided informed consent. The study is reported in accordance with the STROBE statement.^11^

### Inclusion criteria

Consecutive patients aged ≥18 years with chronic gastroduodenal symptoms and negative upper gastrointestinal endoscopy undergoing GEBT were invited to participate. Exclusion criteria included those with known structural gastrointestinal diseases and previous abdominal surgery. Patients with cyclical vomiting syndrome or cannabinoid hyperemesis were also excluded. Specific exclusion criteria related to Gastric Alimetry were a BMI of >35, active abdominal wounds or abrasions, fragile skin, and allergies to adhesives.

### Gastric emptying breath testing

Solid gastric emptying was measured using a 4-hour ^13^C octanoic acid emptying breath test.^12–14^ All subjects were fasted overnight for at least 8 hours ahead of GEBT. Patients were asked to stop medications affecting gastric emptying, such as opioids, prokinetics, anticholinergics, and/or calcium channel blockers at least two days ahead of the GEBT. The test meal used for GEBT was either a pancake with 180 ml of water (11.2 g fat, 31.7 g carbohydrate, 8.4 g protein; 261 kcal total) or an egg with two slices of white toast and 180 ml of water (9.4 g fat, 34 g carbohydrate, 11.5 g protein; 268 kcal total).^12^ Breath samples were taken before starting the test meal and at 15 min intervals for 4 h. The gastric half emptying time (T_1/2_) was calculated as previously described.^15^ Delayed gastric emptying was defined as T_1/2_ >109 min for solids.

### Body surface gastric mapping and symptom profiling

BSGM was performed using the Gastric Alimetry system, which includes a high-resolution stretchable electrode array (8×8 electrodes; 20 mm inter-electrode spacing; 196 cm^2^), a wearable Reader, an iPadOS App and concurrent validated symptom logging during the test.^16–18^ Array placement was preceded by shaving if necessary, and skin preparation (NuPrep; Weaver & Co, CO, USA). Recordings were performed simultaneously with GEBT encompassing 30 min fasting baseline, 10 min meal, and 4 h postprandial recording. Participants are asked to sit reclined in a chair and were asked to limit movement, talking, and sleeping, but were able to read, watch media, work on a mobile device, and mobilize for comfort breaks, although some movement was accepted to deliver breath samples at 15 min intervals in this protocol. Symptom capture included early satiation after meal completion, and symptoms of nausea, bloating, upper gut pain, heartburn, stomach burn, and excessive fullness were measured during continuously testing at 15-minute intervals using 0-10 visual analog scales (0 indicating no symptoms; 10 indicating the worst imaginable extent of symptoms) and combined to form a ‘Total Symptom Burden Score’.^18^

### Metric processing and Interpretation

Standardized metrics were analyzed for both tests.^19,20^ GEBT was assessed using T_1/2_ emptying time, with delay considered T_1/2_ >109 min. BSGM spectral analysis included Principal Gastric Frequency (PGF; reference intervals: 2.65 - 3.35 cycles per minute), BMI-adjusted amplitude (reference intervals: 22 - 70 µV), and Gastric Alimetry Rhythm Index (GA-RI; reference intervals: ≥ 0.25) for BSGM. In addition, a novel BSGM metric was introduced for this study called ‘Meal Response Ratio’ (MRR) to assess meal response timing, calculated as the ratio of the average amplitude in the first 2 hours postprandially to that of the last 2 hours. MRR was not calculated if postprandial recording duration was <4 h. A normal MRR was empirically defined as >1 based on previous studies,^6,19,21^ meaning that the dominant gastric motor response occurred within the first two hours after a meal.

### Statistical analysis

All analyses were performed in Python v3.9.7 and R v.4.0.3 (R Foundation for Statistical Computing, Vienna, Austria). Numerical data were summarized as mean (standard deviation) or median (interquartile range) based on visual and statistical evaluation for normality, with appropriate tests for parametric or non-parametric data performed. Categorical data were cross-tabulated, and differences tested using χ2 or Fisher’s exact tests. Bonferroni corrections were applied for post-hoc corrections.

## Results

Overall, 151 consecutive subjects (118, [78.1%] females, median age 31 [range 18-80] years, BMI median 22 [18.5-35] kg/m^2^) were enrolled and underwent simultaneous BSGM and GEBT. Complete data was available for 143 subjects after excluding 8 (5%) participants due to inadequate test quality. Most patients (87%) successfully completed 100% of the test meal (mean 96±14% meal completion).

Overall (n = 143), the median T_1/2_ was 85 minutes (IQR 31-373), with 25.2% (n = 36/143) classified as having delayed gastric emptying on GEBT. On BSGM testing, 28 (19.6%) had a low GA-RI, 23 (16.1%) had a low BMI-adjusted amplitude, 1 (0.7%) had a low Principal Gastric Frequency, and 12 (8.4%) had a high Principal Gastric Frequency. The novel MRR metric was applied to those with normal spectrograms (n = 93); median MRR was 1.21 (IQR 0.58 - 4.21) with 21 (22.6%) participants classified as having a lagged meal response (i.e. greater gastric amplitude across the latter 2 hours of testing vs first 2 hours of the postprandial period). Those with this lagged meal response phenotype on BSGM had a significantly longer T_1/2_ on GEBT (median 95.0 [IQR 59-373] vs median 78.0 [IQR 31-288], p=0.009; **Figure 1**) and a higher rate of delayed emptying (42.9% [9/21] vs 16.7% [12/72], p = 0.03).

**Figure 1:**
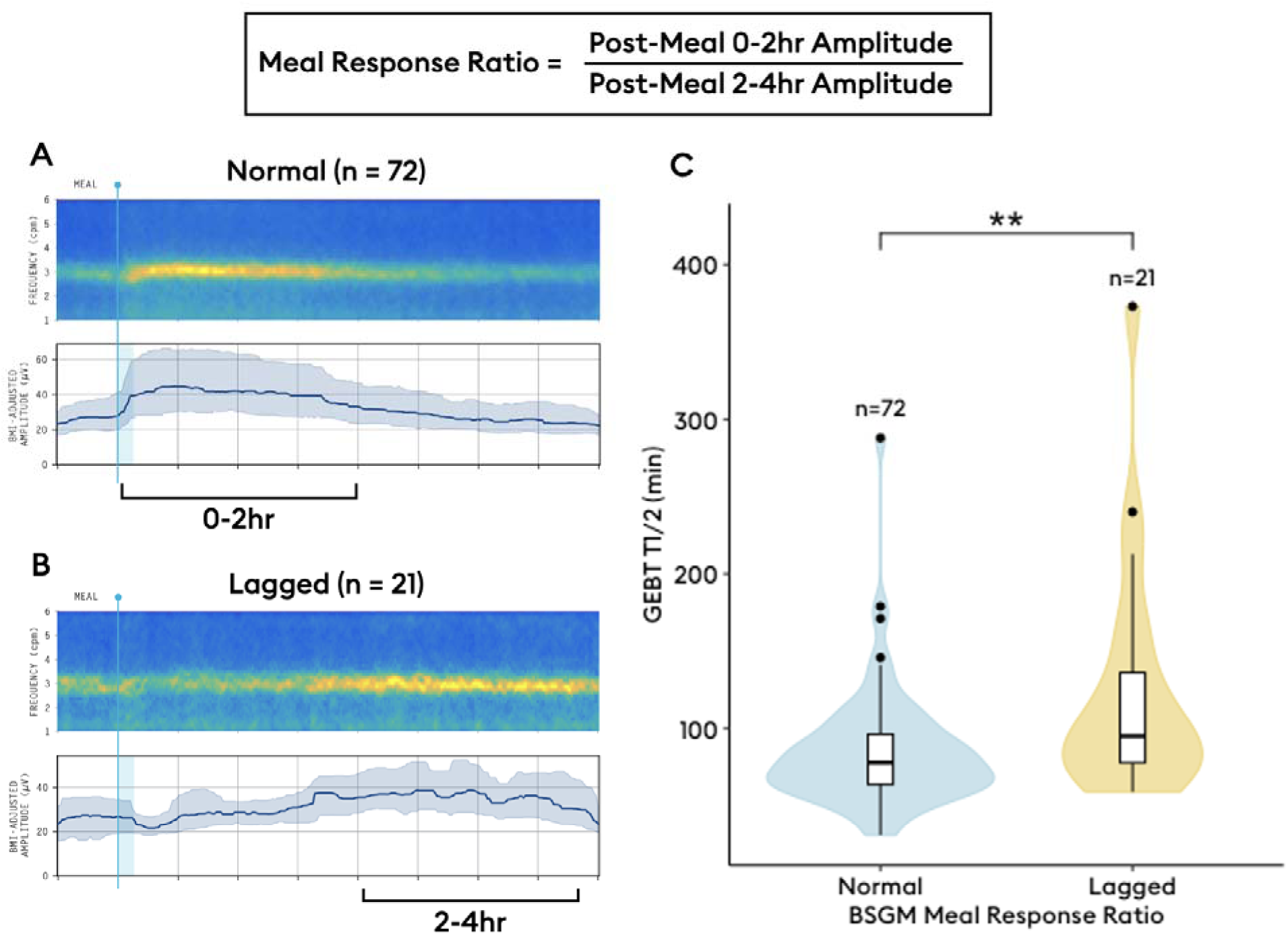
A) Average spectrogram of patients with normal BSGM meal response (n=72). B) Average spectrogram of patients with lagged BSGM meal response (n=21). C) Box plots showing higher rates of delayed gastric emptying for patients with lagged BSGM meal response (p=0.002).

Among those with delayed gastric emptying on GEBT (n = 36/143, 25.2%), the following BSGM phenotypes were identified: 12 (33.3%) had a normal spectral analysis, 9 (25.0%) had a lagged meal response phenotype (MRR ≤1), 11 (30.6%) had a low amplitude or GA-RI, and 4 (11.1%) had a high PGF (**Figure 2**). When emptying was normal (n = 107/143, 74.8%), 28 (26.2%) had a low amplitude or GA-RI, 7 (6.5%) had a high PGF, 12 (11.2%) had a lagged meal response phenotype, and 60 (56.1%) had a normal BSGM. Notably the lagged meal response occurred more frequently in those with delayed emptying, (42.9% [9/21] vs 16.7% [12/72], p = 0.03). Frequency of each phenotype by gastric emptying status is shown in **Figure 3**.

**Figure 2:**
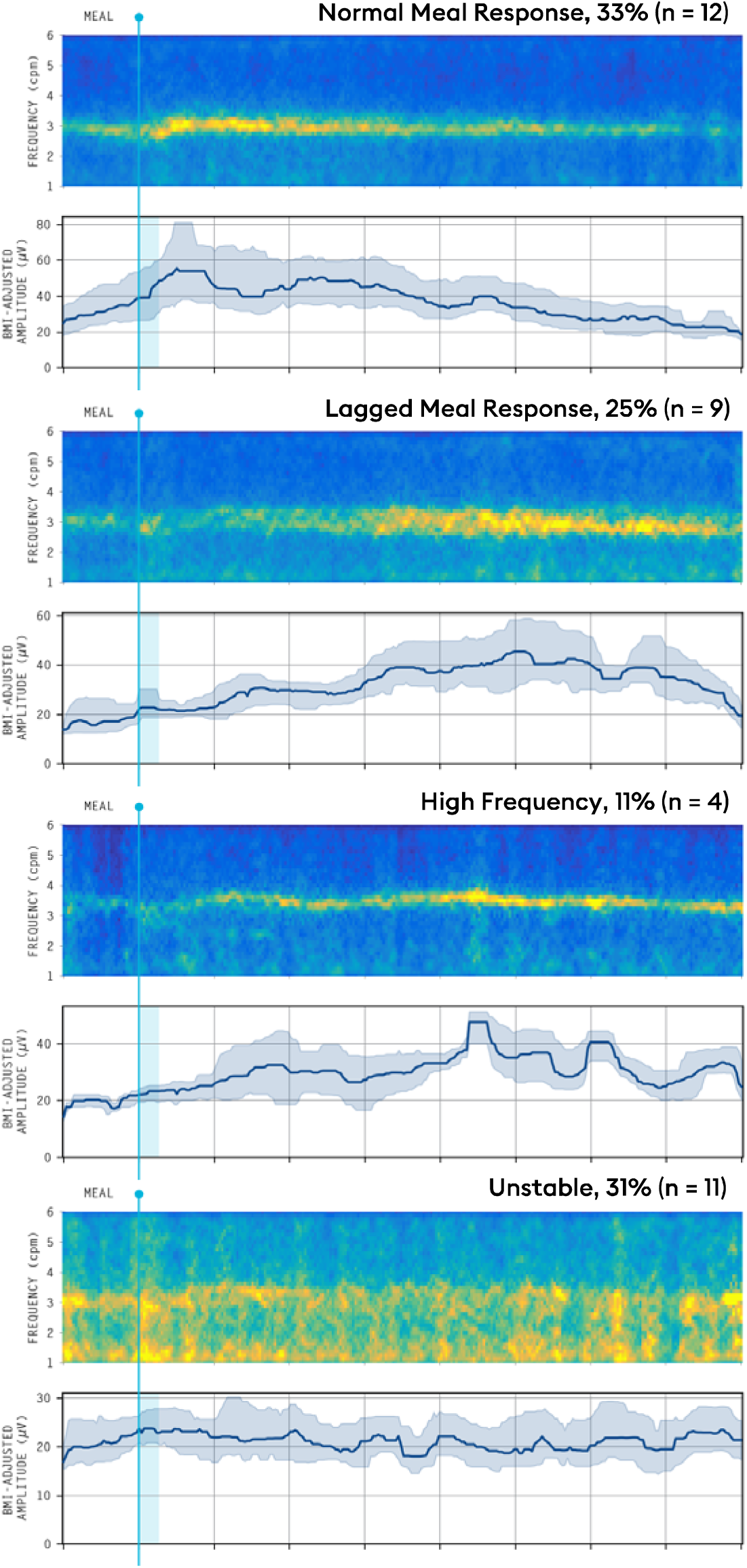
Phenotypes of delayed gastric emptying. Contributing mechanisms for delayed gastric emptying reported for the 36 (25.2%) of patients with delayed gastric emptying on gastric emptying breath testing.

**Figure 3:**
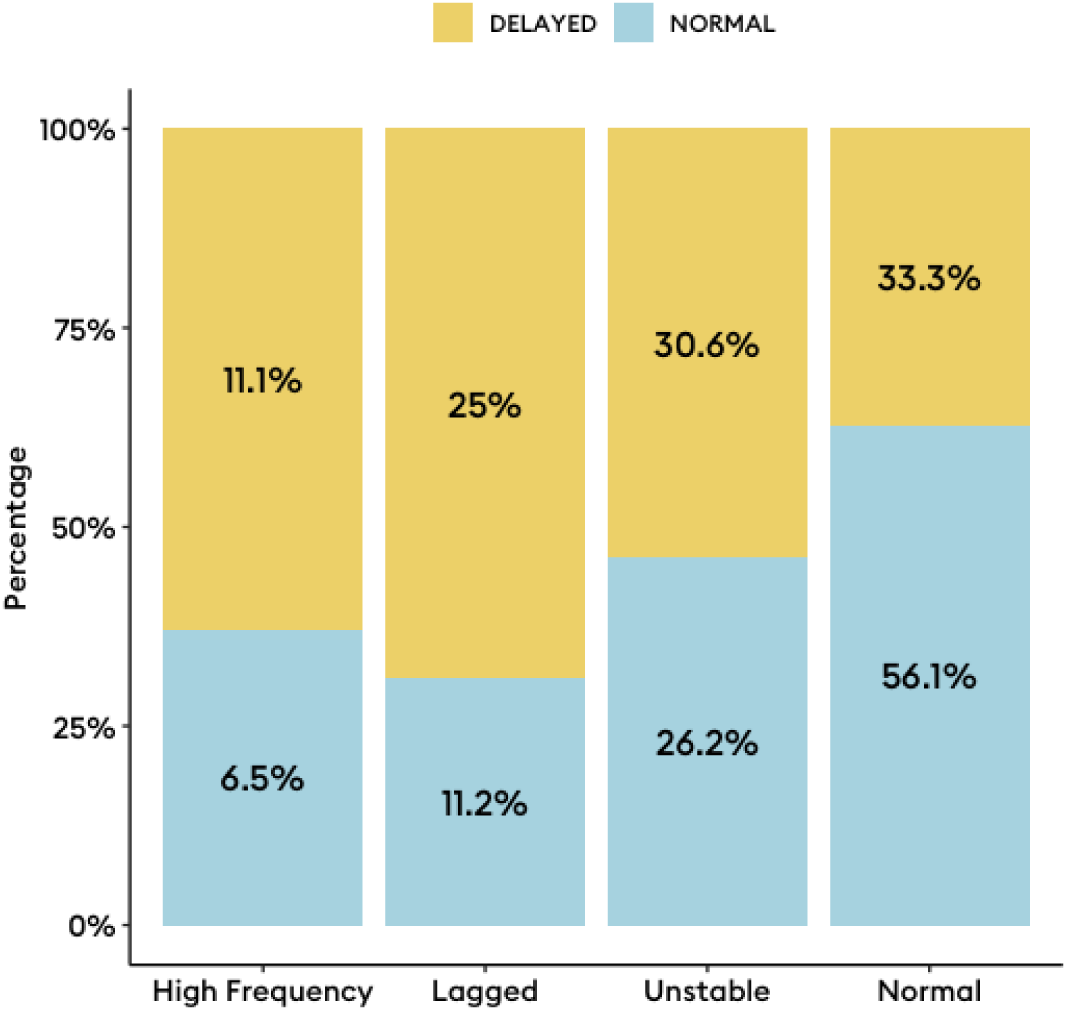
Proportion of each body surface gastric mapping phenotype with delayed and normal gastric emptying breath test results. Percentages reflect the proportion of each phenotype within their respective emptying classification.

Symptom comparisons across the whole cohort showed no differences in any symptoms between BSGM phenotypes (**Figure 4**; all comparisons p>0.05). Participants with delayed gastric emptying had worse symptoms (**Figure 4B**, p = 0.003), with significant differences observed for nausea, upper gut pain, excessive fullness, and early satiety (**Table 1**). However, correlations between delayed transit and ‘Total Symptom Burden Score’ were weak (r = 0.18, p = 0.03). There were no other differences in symptom severity between those with delayed and normal emptying across phenotypes (**Table S1**; all comparisons p>0.05). In those with delayed emptying, there was a mean 2-point lower nausea score when a lagged meal response was present (p = 0.049).

**Figure 4:**
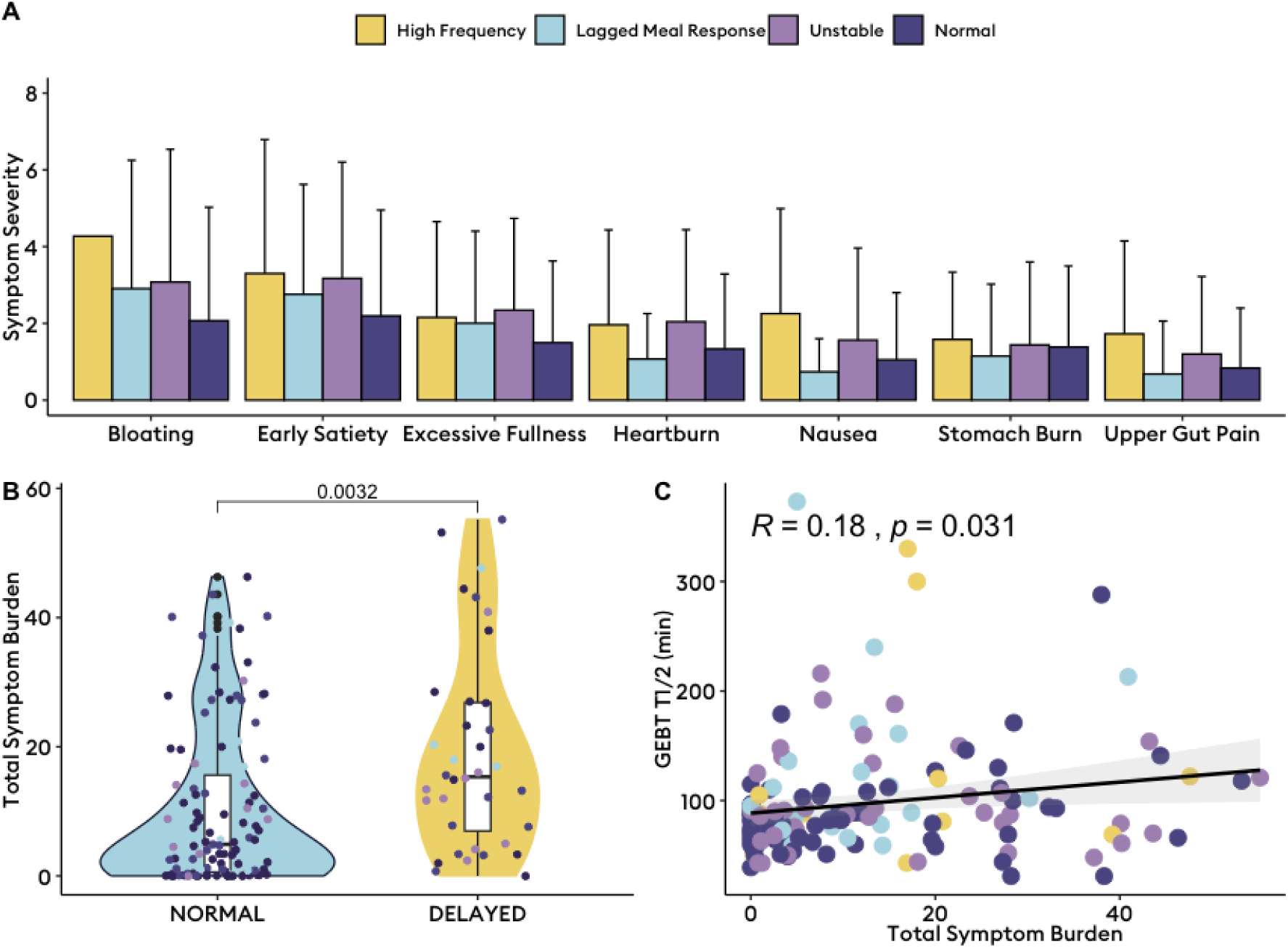
Symptom variation across body surface gastric mapping (BSGM) phenotypes. A) Mean and upper boundary of the standard deviation plotted across each symptom stratified by BSGM phenotype. There were no statistically significant differences in symptom severity across body surface gastric mapping phenotypes (p>0.05). B) Total symptom burden (0 - 70) between those with delayed and normal gastric emptying on gastric emptying breath testing (GEBT). C) A weak correlation between slower gastric emptying as measured by the T_1/2_ on GEBT was shown with dots colored by BSGM phenotype.

**Table 1:** Time-of-test symptom severity by delayed gastric emptying status based on gastric emptying breath testing after post-hoc correction.

| Symptom | Delayed GEBT | Normal GEBT | p |
| --- | --- | --- | --- |
| Nausea | 2.4 (2.6) | 1.0 (1.7) | 0.001 |
| Bloating | 2.8 (2.6) | 1.5 (2.1) | 0.004 |
| Upper Gut Pain | 2.4 (2.3) | 1.2 (1.9) | 0.004 |
| Heartburn | 1.2 (2.0) | 0.9 (1.7) | 0.325 |
| Stomach Burn | 1.6 (2.2) | 1.1 (1.9) | 0.202 |
| Excessive Fullness | 4.2 (3.2) | 2.1 (2.6) | <0.001 |
| Early Satiety | 4.3 (3.5) | 2.1 (3.0) | <0.001 |

*GEBT, gastric emptying breath testing.*

## Discussion

This study aimed to define specific gastroparesis subgroups based on simultaneous BSGM and GEBT testing. We also introduced a new ‘meal response ratio’ (MRR) metric to quantify the dynamic post-prandial motor function of the stomach, and found a lagged meal response (MRR ≤1) was correlated with delayed emptying. Using BSGM metrics of gastric function, four specific subgroups of gastroparesis were identified: firstly, a normal spectrogram group with appropriately timed postprandial gastric motor activity (33%); secondly, a lagged meal response with a delayed onset to gastric motor activity (25%); thirdly, an unstable spectrogram group with low rhythm stability (31%); finally, an elevated gastric frequency group (11%). Whereas symptoms alone fail to separate mechanistic groups, the addition of BSGM testing allowed mechanistic phenotyping with potential to facilitate targeted disease management.

There has been recent controversy around the diagnostic value of gastric emptying testing.^3^ Gastric emptying scintigraphy testing alters clinical management in <50% of cases,^22^ and clinicians are often required to make treatment decisions based on symptoms alone. It is well established that symptoms poorly differentiate chronic gastroduodenal disorders,^3,7^ owing to significant overlap between diagnostic categories^23^ and multiple disease mechanisms contributing to individual symptoms. Given symptoms and transit testing have pitfalls in informing management in gastroparesis, more specific tests of gastric function referring to underlying pathophysiology are desirable.^10^ Gastric Alimetry has been shown to phenotype patient subgroups and direct care in chronic gastroduodenal disorders,^8,21,24^ and therefore we applied Gastric Alimetry testing here to reveal novel subtypes of gastroparesis based on mechanisms for delayed gastric emptying which could provide therapeutic targets.

Gastric transit is a higher order function that can result from several possible derangements of gastric function. Antral hypomotility may arise secondary to discoordinated motor activity and/or damage to ICC networks, as has been shown in patients with gastroparesis with dysrhythmic myoelectrical activity.^25,26^ In addition, autonomic dysfunction has been separately implicated in impaired accommodation and delayed emptying.^27,28^ Reduced accommodation may be evidenced by low intragastric meal distribution (i.e., antral retention), which has been correlated with symptom burden in gastroparesis.^29^ Decreased gastric tone may additionally result in inadequate gastroduodenal pressure gradients to facilitate transit,^30,31^ possibly also contributing to delayed emptying through fundic retention.^30^ Finally, pylorospasm or increased pyloric tone, could also be contributory as suggested by favorable results of endoscopic pyloromyotomy in patients with refractory gastroparesis.^32–34^

The phenotypes identified with the aid of BSGM in this study likely relate to the various underlying gastroparesis pathophysiologies discussed above. This includes characterization of those patients with a neuromuscular phenotype through a low GA-RI,^21,35^ and those with vagal neuropathies through an elevated PGF.^36,37^ In addition, we postulate the MRR could offer a novel way to differentiate proximal and distal gastric causes to delayed transit (**Supplementary Figure 1**). For example, when MRR ≤1 implies a relative delay to onset of gastric activity in excess of 2 h, which may relate to reduced postprandial gastric tone. This pattern is frequently resulting in symptoms correlating to the lagged meal response period, with symptoms then improving following the onset of gastric activity.^7^ Alternatively, when MRR is >1 but transit is delayed, this suggests an intact neuromuscular apparatus likely generating effective antral contractions, plausibly implicating antropyloric discoordination or a functional pyloric obstruction, which has previously been shown in the electrogastrography literature in association with sustained myoelectrical amplitudes.^32^

Several limitations of this study are noted. The use of a non-standard meal for Gastric Alimetry testing has been associated with lower amplitudes and GA-RI than the standard meal, but all metrics have been observed to be consistent between pancake and egg meals.^38^ In addition, while these concepts outlined above propose to advance the mechanistic understanding of the subgroups introduced in this study based on known physiology, validation is required. This could include reference against additional modalities such as scintigraphic intragastric meal distribution, antral contractility and motility indices;^39,40^ single photon emission computed tomography (SPECT); or dynamic MRI. Ongoing efforts for simultaneous validation with these modalities against BSGM are now underway. If these hypotheses are confirmed, they could enable targeted therapies for different subgroups of patients with gastroparesis, which could be evaluated in outcomes studies. Candidate agents include buspirone for impaired accommodation,^41^ or gastric per oral endoscopic myotomy for pylorus-related causes for gastroparesis.^33,42^

Future work could also specifically evaluate whether these proposed mechanism match those presented in a comprehensive study of 1287 patients with chronic gastroduodenal symptoms, in which SPECT was used to assess fundic accommodation and gastric emptying scintigraphy was used to assess gastric emptying.^31^ In this previous study, 21.1% of patients had abnormal accommodation and emptying, potentially correlating with our cohort showing delayed GEBT and lagged meal response phenotype. A further 29.8% had normal emptying and accommodation, potentially correlating with our cohort showing normal BSGM and normal GEBT. 21.9% had abnormal accommodation with normal emptying, which could correspond to our cohort with normal emptying and a long lag phenotype and/or high PGF. Finally, 27.1% of patients had delayed emptying alone which could reflect our cohort with delayed GEBT and normal BSGM.

Interestingly, delayed emptying was associated with increased symptoms in this study, which has not been a consistent finding of previous gastric emptying literature.^4,43,44^ It is plausible that the positive symptom associations reflect the advantages of robust and validated time-of-test symptom capture using a symptom logging App incorporating pictograms^18^, compared to alternative symptom assessments relying on recall over previous weeks, which have inconsistently detected this relationship elsewhere.^4^

In summary, this study presents mechanism-based phenotypes of gastroparesis based on simultaneous BSGM and GEBT. We quantify dynamic gastric activity through the ‘MRR’, a novel biomarker for delayed gastric motor activity. BSGM extends the characterization of gastric sensorimotor function, towards the goal of achieving more specific diagnostic phenotypes to direct targeted therapies.

## Data Availability

All data produced are available online at

## Acknowledgements

N/A

---

**Table S1:** Post-hoc comparisons of symptom severity by gastric emptying breath test status among each body surface gastric mapping phenotype.

| Phenotype | Symptom | GEBT Status |  | Delayed GEBT (n) | Normal GEBT (n) | Mean Difference | p | Adjusted p value | Significance |
| --- | --- | --- | --- | --- | --- | --- | --- | --- | --- |
| High Frequency | Bloating | Delayed | Normal | 4 | 7 | 1.24 | 0.27 | 1 | ns |
| Delayed Meal Response | Bloating | Delayed | Normal | 9 | 12 | 0.989 | 0.34 | 1 | ns |
| Unstable | Bloating | Delayed | Normal | 11 | 28 | 0.639 | 0.531 | 1 | ns |
| Normal | Bloating | Delayed | Normal | 12 | 60 | 1.77 | 0.0994 | 1 | ns |
| High Frequency | Early Satiety | Delayed | Normal | 4 | 7 | 0.86 | 0.414 | 1 | ns |
| Delayed Meal Response | Early Satiety | Delayed | Normal | 9 | 12 | 1.701 | 0.111 | 1 | ns |
| Unstable | Early Satiety | Delayed | Normal | 11 | 28 | -0.083 | 0.935 | 1 | ns |
| Normal | Early Satiety | Delayed | Normal | 12 | 60 | 3.49 | 0.00372 | 0.10416 | ns |
| High Frequency | Excessive Fullness | Delayed | Normal | 4 | 7 | 2.561 | 0.047 | 1 | ns |
| Delayed Meal Response | Excessive Fullness | Delayed | Normal | 9 | 12 | 0.132 | 0.896 | 1 | ns |
| Unstable | Excessive Fullness | Delayed | Normal | 11 | 28 | 0.87 | 0.394 | 1 | ns |
| Normal | Excessive Fullness | Delayed | Normal | 12 | 60 | 2.99 | 0.0105 | 0.294 | ns |
| High Frequency | Heartburn | Delayed | Normal | 4 | 7 | 0.553 | 0.606 | 1 | ns |
| Delayed Meal Response | Heartburn | Delayed | Normal | 9 | 12 | -0.816 | 0.427 | 1 | ns |
| Unstable | Heartburn | Delayed | Normal | 11 | 28 | 0.171 | 0.866 | 1 | ns |
| Normal | Heartburn | Delayed | Normal | 12 | 60 | 1.195 | 0.252 | 1 | ns |
| High Frequency | Nausea | Delayed | Normal | 4 | 7 | 1.74 | 0.169 | 1 | ns |
| Delayed Meal Response | Nausea | Delayed | Normal | 9 | 12 | 0.414 | 0.687 | 1 | ns |
| Unstable | Nausea | Delayed | Normal | 11 | 28 | 0.616 | 0.548 | 1 | ns |
| Normal | Nausea | Delayed | Normal | 12 | 60 | 3.323 | 0.00532 | 0.14896 | ns |
| High Frequency | Stomach Burn | Delayed | Normal | 4 | 7 | 0.039 | 0.97 | 1 | ns |
| Delayed Meal Response | Stomach Burn | Delayed | Normal | 9 | 12 | 0.846 | 0.412 | 1 | ns |
| Unstable | Stomach Burn | Delayed | Normal | 11 | 28 | -0.483 | 0.635 | 1 | ns |
| Normal | Stomach Burn | Delayed | Normal | 12 | 60 | 1.885 | 0.0823 | 1 | ns |
| High Frequency | Upper Gut Pain | Delayed | Normal | 4 | 7 | 1.53 | 0.183 | 1 | ns |
| Delayed Meal Response | Upper Gut Pain | Delayed | Normal | 9 | 12 | -0.129 | 0.899 | 1 | ns |
| Unstable | Upper Gut Pain | Delayed | Normal | 11 | 28 | 0.906 | 0.377 | 1 | ns |
| Normal | Upper Gut Pain | Delayed | Normal | 12 | 60 | 2.348 | 0.0355 | 0.994 | ns |

**Supplementary Figure 1:**
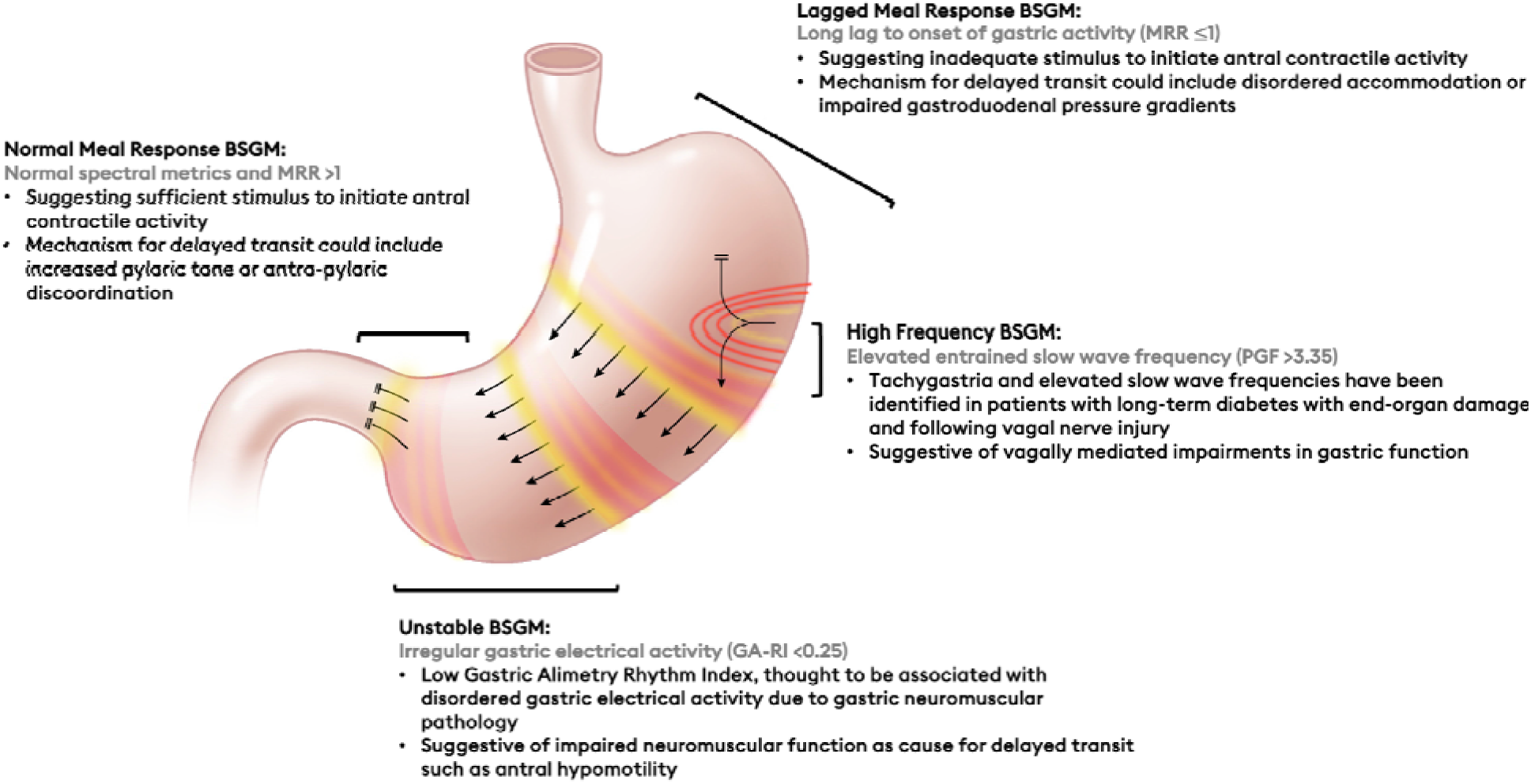
Putative mechanisms for gastroparesis mapped to each body surface gastric mapping phenotype. Figure adapted and customized with permission from O’Grady et al.^45^

